# Co-circulation of two lineages of Oropouche virus in the Amazon basin, Colombia, 2024

**DOI:** 10.1101/2024.09.04.24312892

**Authors:** Jaime Usuga, Daniel Limonta, Laura S. Perez-Restrepo, Karl A. Ciuoderis, Isabel Moreno, Angela Arevalo, Vanessa Vargas, Michael G. Berg, Gavin A. Cloherty, Juan P. Hernandez-Ortiz, Jorge E. Osorio

**Affiliations:** School of Veterinary Medicine, University of Wisconsin, Madison, Wisconsin, USA; Global Health Institute One Health Colombia, National University of Colombia, Medellin, Colombia; Infectious Diseases Research, Abbott Diagnostics and Abbott Pandemic Defense Coalition, Abbott Park, Illinois, USA; National University of Colombia, Medellín, Colombia; Global Health Institute, University of Wisconsin, Madison, Wisconsin, USA

**Keywords:** Oropouche, Colombia, reassortment, fever, sequencing, outbreak, Leticia, lineages, Amazon

## Abstract

In early 2024, explosive outbreaks of Oropouche virus (OROV) linked to a novel viral lineage were documented in the Brazilian Amazon. Here, we report the introduction of this emerging orthobunyavirus into Colombia and its co-circulation with another OROV lineage. This investigation highlights the complex arbovirus dynamics in South America.

---

Oropouche virus (OROV) is a reemerging arbovirus belonging to the *Orthobunyavirus* genus, *orthobunyavirus oropoucheense* species, of the family *Peribunyaviridae*. The three single-stranded, negative-sense RNA segments (L: Large, M: Medium, S: Small) of the OROV genome allow reassortments (1). Besides acute fever, OROV can cause meningitis and encephalitis. While biting midges, *Culicoides paraensis*, are the main vector of OROV in urban cycles, different insect species are vectors in sylvatic cycles. Vertebrates, such as primates, small mammals, and possibly birds, serve as reservoir hosts (1,2).

OROV has spread rapidly in South America causing over 500,000 infections (2,3). Most OROV cases have occurred in the Brazilian Amazon, where explosive outbreaks affecting sometimes thousands of inhabitants, have been reported after OROV identification in Trinidad and Tobago in 1955 (2,4,5). In a study conducted by our group in 2019-2021 in Colombia, we found that OROV and dengue virus (DENV) were responsible for over 10 and 20% of acute febrile illness, respectively, at four sites (Cúcuta, Cali, Villavicencio, and Leticia). The phylogenetic analysis revealed two separate OROV introductions into Colombia from bordering Ecuador (Cúcuta, Cali, and Villavicencio) and Peru (Leticia) (6).

In early February 2024, the PAHO/WHO issued an epidemiological alert because of the dramatic increase in OROV cases in four Amazonian states of Brazil (7). Over 1000 confirmed cases by RT-qPCR have been reported in the state of Amazonas alone between 2023 and January 2024 (8). Brazilian authors recently shared in a pre-publication the sequence data of the OROV causing this large-scale outbreak. After analyzing over 100 full-length genomes, they identified multiple reassortment events and concluded that this virus is a new lineage (OROV_BR-2015-2023_) (9). In this work, we are reporting the first OROV cases in Colombia caused by this newly identified viral lineage, which is co-circulating with another previously characterized OROV lineage (6).

## The Study

In early January 2024, health authorities noticed a slight increase in acute febrile cases in the Leticia municipality, Amazonas department of Colombia, which has over 53,000 inhabitants. One-hundred and seventeen individuals complaining of chills (94.6%), headache (87.2%), arthralgia (65%), myalgia (41%), diarrhea (34.2%), fatigue (33.3%), and rash (6%) were treated at the emergency room of the Leticia hospital over five weeks. While fever (≥38.5°C) was confirmed at the doctor’s office in >46% of patients, less than 3% presented respiratory symptoms. No hemorrhagic manifestations were present in the febrile patients, nor were severe illnesses or hospitalizations documented among the studied individuals. Cases were between 7 and 92-years -old. Sixty individuals were male, and fifty-seven were female. After obtaining the written consent from adults and children’s parents (or legal guardians), serum samples were collected. The protocol for this study was approved by the ethics committee of the Corporacion de Investigaciones Biologicas (SC-6230-1).

Frozen serum samples were shipped by air over 1300 km to One Health Colombia, located in Medellin city. One Health Colombia is a center established between the Global Health Institute at the University of Wisconsin-Madison and the National University of Colombia-Medellin. Following the center guidelines at One Health Colombia (6), serum samples were tested for arboviruses (zika virus or ZIKV, mayaro virus or MAYV, chikungunya virus or CHIKV, DENV, OROV), hepatitis B and C viruses, *Leptospira* spp., and *Plasmodium* spp. using published and Abbott (Illinois, US) protocols (see **Appendix** and references therein).

DENV genome was detected by ZCD (zika, chikungunya, dengue) Trioplex real-time RT-qPCR in eight samples, which were also DENV non-structural protein 1 positive (N=4) or anti-DENV IgM positive (N=3). The study population had a high anti-DENV IgG prevalence of 77.7% (N=117). All samples were negative for ZIKV, CHIKV, and MAYV RNA. Seven samples presented *Plasmodium* spp. DNA by qPCR, with five samples showing positivity for the *P. vivax* antigen. No samples were positive for hepatitis B virus surface Ag, anti-hepatitis C virus antibody, and *Leptospira* spp. DNA.

Using our recently designed OROV RT-qPCR assay (6), OROV RNA of L and M segments were detected in eight samples. Next, NGS sequencing was conducted using metagenomic and target enrichment approaches in these eight serum specimens, as described previously (6). Phylogenetic analysis using available OROV genomes and the sequences of the new Brazilian clade, OROV_BR-2015-2023_ (9,10), revealed its presence in two samples from Leticia, LET-2099 and LET-2102 (**Figure 1**). While the L and S segments each branch as paraphyletic clades basal to OROV_PE/CO/EC-2008-2021_, its M segment is more closely related to OROV_BR-2009-2018_ sequences. Amino acid changes of this new lineage compared to ancestral viral strains indicated multiple mutations in L and M segments (**Appendix**). In addition, six samples were detected from the OROV_PE-CO-EC/2008-2021_ lineage recently reported as circulating in 2019-2021 in Colombia (6) (**Table**). OROV cases did not exhibit co-infections aside from clinical manifestations of acute undifferentiated fever **(Appendix**) and they had not traveled outside of the Leticia municipality and its surrounding areas in the previous two weeks (**Figure 2**). Six full-length genomes of OROV analyzed in this work, including the two novel OROV, were deposited in the GenBank under accession numbers PP477303-PP477320.

**Figure 1.**
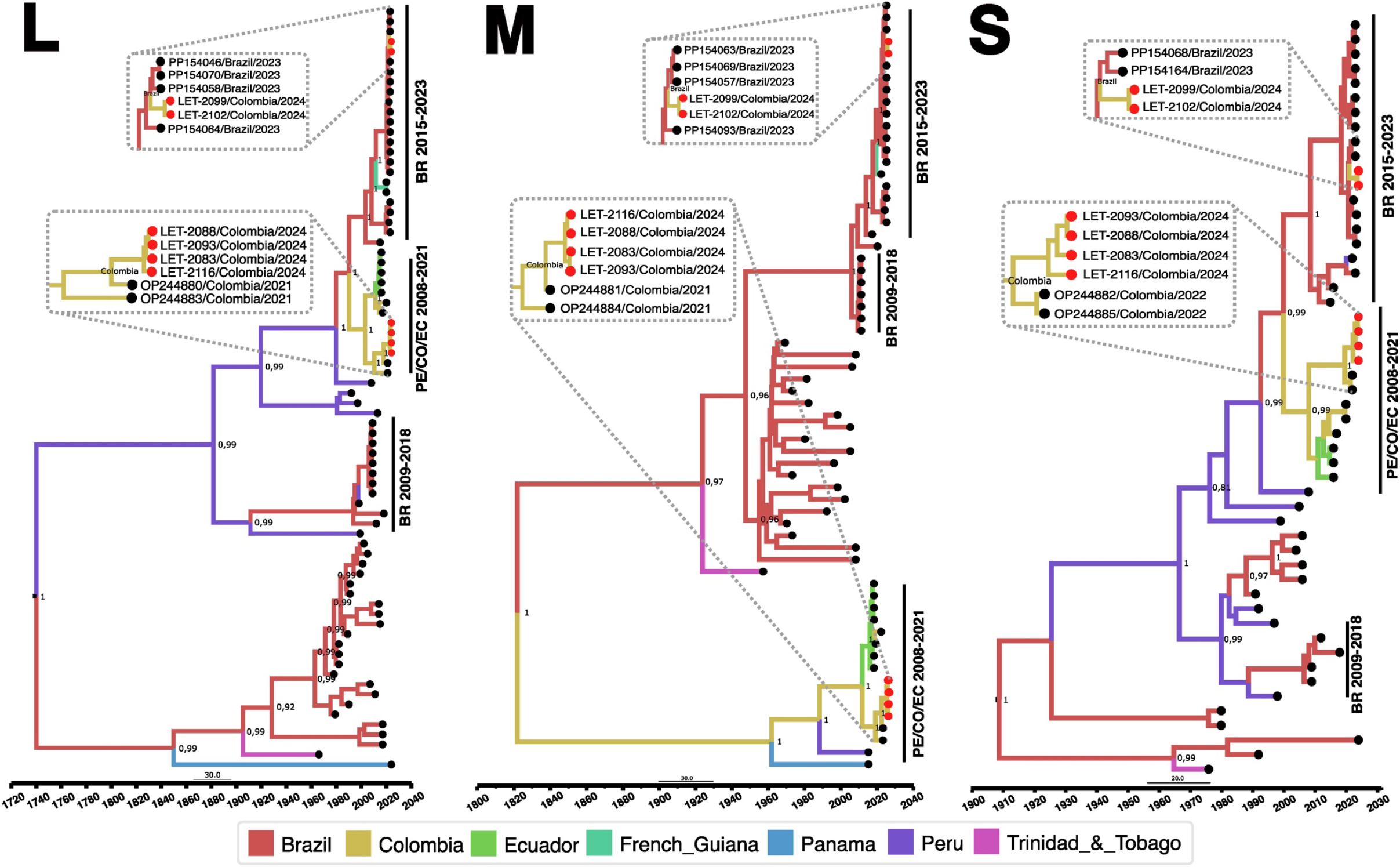
Time-scaled Bayesian phylogeographic tree of Oropouche viruses from Colombia. Bayesian phylogenetic trees of L (left), M (center) and S (right) gene segments were estimated using Beast (Bayesian Markov-Chain Monte Carlo > 100,000,000 generations) and IQ-Tree in ModelFinder (ultrafast bootstrapping and 1,000 replicates). Best-fit model was selected according to Bayesian information criteria. Strict molecular clock model was used for L and S segments, while an uncorrelated relaxed molecular clock model was used for the M segment. The phylogeny branches are colored according to their descendant place of origin (figure key). Bayesian posterior values (≥0.8) are annotated at specific nodes of the trees. Sequences from this study were compared to reference sequences from other studies. Main clusters are denoted with the following reference labels: BR 2015-2023 cluster represents the recent outbreak of the new lineage of OROV in Brazil from 2015 to 2023; PE/CO/EC 2008-2021 cluster represents sequences from Colombia, Peru, and Ecuador from 2008 to 2021; and BR 2009-2018 cluster represents sequences from Brazil from 2009 to 2018. Viruses from this study are represented by red circles with Let as the abbreviation of Leticia.

**Figure 2.**
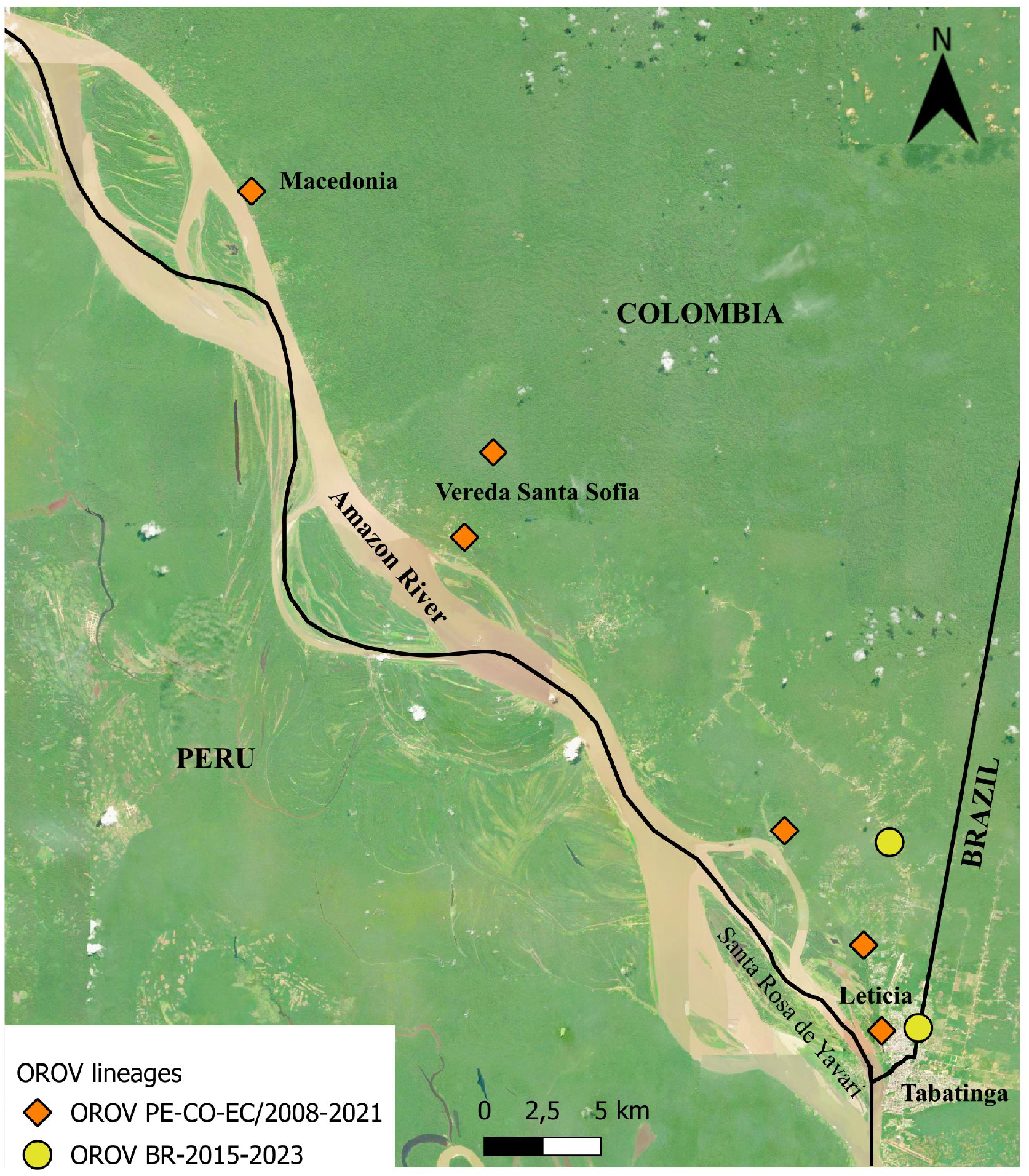
Map of the Leticia municipality and the Three Borders (Colombia, Peru, and Brazil) in the Amazon basin, Colombia. The map shows where the eight cases of OROV reside in the Leticia municipality. Yellow dots indicate cases with the newly identified OROV_BR-2015-2023_. Macedonia, Vereda Santa Sofia, Santa Rosa de Yavari, and Tabatinga correspond to indigenous communities or cities. Map generated using QGIS Version 3.36.0 RC (03/12/2024).

**Table 1.** Case identification, demographic information, blood sampling date, and OROV clades of the eight febrile cases with OROV in Leticia, Colombia, 2024.

| ID* | Sex | Sampling<br>date | Ct value | OROV† clade |
| --- | --- | --- | --- | --- |
| LET‡-2040 | Male | 2024 Jan | 32.9 | OROV<sub>PE-CO-EC/2008-2021</sub> |
| LET-2083 | Male | 2024 Feb | 28.6 | OROV<sub>PE-CO-EC/2008-2021</sub> |
| LET-2088 | Male | 2024 Feb | 31.5 | OROV<sub>PE-CO-EC/2008-2021</sub> |
| LET-2093 | Female | 2024 Feb | 33.2 | OROV<sub>PE-CO-EC/2008-2021</sub> |
| LET-2099 | Male | 2024 Feb | 28.7 | OROV<sub>BR-2015-2023</sub> |
| LET-2102 | Male | 2024 Feb | 30.1 | OROV<sub>BR-2015-2023</sub> |
| LET-2116 | Female | 2024 Feb | 34.2 | OROV<sub>PE-CO-EC/2008-2021</sub> |
| LET-2117 | Female | 2024 Feb | 34.4 | OROV<sub>PE-CO-EC/2008-2021</sub> |

\*ID, identification.
†OROV, Oropouche virus.
‡Let, Leticia.

## Conclusions

In the present cross-sectional study, we are reporting for the first time the circulation in Colombia of OROV_BR-2015-2023_, a novel orthobunyavirus that was very recently documented in Brazil (9,10). Thus, this is the first report of the newly identified lineage of OROV outside Brazil. Given the numerous mutations in RdRp and glycoproteins for OROV _BR-2015-2023_, enhanced replication and immune evasion capabilities likely increase viral fitness and transmission. In addition, we detected in six patients the co-circulation of an OROV lineage identified previously by our group in Colombia (6).

The identification of co-circulating strains of OROV exemplifies the evolving nature of orthobunyaviruses and raises concerns about future reassortment events and the emergence of new lineages with more severe clinical phenotypes and enhanced vector competence. In an arbovirus hotspot such as Leticia, it is not known if cross-protective immunity will exist between these two OROV lineages. Previously, a reassortant OROV isolated from outbreaks in Iquitos, the largest city in the Peruvian Amazon, provided limited cross-protection against an OROV strain (11).

Leticia municipality, the southernmost region in Colombia (**Figure 2**), remains isolated from the rest of Colombia’s road network, and thus travelers typically reach Leticia by air. The capital of the Colombian Amazonas department, Leticia city, is in Leticia municipality where the borders of Colombia, Brazil, and Peru converge, forming an area known as the Three Borders. Tabatinga city, in the Brazilian state of Amazonas, is located right across from Leticia city forming a unique suburban area nearby the Peruvian city of Santa Rosa de Yavarí, which is on an island in the Amazon River (12). Due to the significant human mobility related to local businesses, it is very likely that the new OROV lineage is spreading rapidly in these borderlands. Furthermore, we believe that the novel OROV in Leticia came from either a nearby settlement or air travelers from the Brazilian Amazon, where persistent outbreaks are occurring.

Our One Health center has previously reported outbreaks of ZIKV (13), DENV (14), OROV (6), and more recently has detected yellow fever virus (unpub. data) and MAYV (15) in Colombia, which underscores the dynamic landscape of reemerging arboviruses in this region. The sustained transmission of established strains and the unpredictable reassortment events of OROV able to cause outbreaks as described here is yet another demonstration of the public health challenges of surveillance, prevention, and control of arbovirus reemergence in the Americas.

## Data Availability

All data produced in the present work are contained in the manuscript

## Acknowledgments

We would like to thank the clinical and laboratory staff of the health-care institutions from Leticia city (Hospital San Rafael de Leticia, and Fundación Clínica Leticia) for their participation in our study. We also thank the staff of One Health Colombia, particularly the laboratory personnel, for their support in this work. Juan P. Hernandez-Ortiz and Laura S. Perez-Restrepo express their gratitude to Fundación Fraternidad Medellin and Fundación Sofia Perez de Soto for their unwavering support for the One Health Ph.D. awards. We are grateful for the map of the Leticia municipality in the Amazon basin, Colombia, generated by Celeny Ortiz Restrepo from One Health Colombia. This research was supported by the Global Health Institute of the University of Wisconsin-Madison, the National University of Colombia-Medellin, and the virus discovery research program of the Abbott Pandemic Defense Coalition (APDC).

## Notes

### Competing Interest Statement

The authors have declared no competing interest.

### Author Declarations

The protocol for this study was approved by the ethics committee of the Corporacion de Investigaciones Biologicas (SC-6230-1) in Colombia

### Summary of Updates

We have added a new reference to the reference list.

## References

1. Wesselmann KM, Postigo-Hidalgo I, Pezzi L, de Oliveira-Filho EF, Fischer C, de Lamballerie X, et al. Emergence of Oropouche fever in Latin America: a narrative review. Lancet Infect Dis. 2024 Jan 25;

2. Pinheiro FP, Travassos da Rosa AP, Travassos da Rosa JF, Ishak R, Freitas RB, Gomes ML, et al. Oropouche virus. I. A review of clinical, epidemiological, and ecological findings. Am J Trop Med Hyg. 1981 Jan;30(1):149–60.

3. Forshey BM, Guevara C, Laguna-Torres VA, Cespedes M, Vargas J, Gianella A, et al. Arboviral etiologies of acute febrile illnesses in Western South America, 2000-2007. PLoS Negl Trop Dis. 2010 Aug 10;4(8):e787.

4. Anderson CR, Spence L, Downs WG, Aitken TH. Oropouche virus: a new human disease agent from Trinidad, West Indies. Am J Trop Med Hyg. 1961 Jul;10:574–8.

5. Azevedo RS da S, Nunes MRT, Chiang JO, Bensabath G, Vasconcelos HB, Pinto AY das N, et al. Reemergence of Oropouche fever, northern Brazil. Emerging Infect Dis. 2007 Jun;13(6):912–5.

6. Ciuoderis KA, Berg MG, Perez LJ, Hadji A, Perez-Restrepo LS, Aristizabal LC, et al. Oropouche virus as an emerging cause of acute febrile illness in Colombia. Emerg Microbes Infect. 2022 Dec;11(1):2645–57.

7. Pan American Health Organization, World Health Organization. Epidemiological Alert: Oropouche in the Region of the Americas. Washington, D.C.; 2024 Feb.

8. Pan American Health Organization, World Health Organization. Public Health Risk Assessment related to Oropouche Virus (OROV) in the Region of the Americas. Washington, D.C.; 2024 Feb.

9. Iani FC de M, Mota Pereira F, de Oliveira EC, Nascimento Rodrigues JT, Hoffmann Machado M, Fonseca V, et al. Rapid spatial Expansion Beyond the Amazon Basin: Oropouche Virus joins other main arboviruses in epidemic activity across the Americas. medRxiv. 2024 Aug 4;

10. Naveca FG, de Almeida TAP, Souza V, Nascimento V, Silva D, Nascimento F, et al. Emergence of a novel reassortant Oropouche virus drives persistent human outbreaks in the Brazilian Amazon region from 2022 to 2024. medRxiv. 2024 Jul 24;

11. Aguilar PV, Barrett AD, Saeed MF, Watts DM, Russell K, Guevara C, et al. Iquitos virus: a novel reassortant Orthobunyavirus associated with human illness in Peru. PLoS Negl Trop Dis. 2011 Sep 20;5(9):e1315.

12. Farnsworth-Alvear A, Palacios M, López AMG, editors. The colombia reader: history, culture, politics. Duke University Press; 2016.

13. Camacho E, Paternina-Gomez M, Blanco PJ, Osorio JE, Aliota MT. Detection of autochthonous zika virus transmission in sincelejo, colombia. Emerging Infect Dis. 2016 May;22(5):927–9.

14. Ciuoderis KA, Usuga J, Moreno I, Perez-Restrepo LS, Flórez DY, Cardona A, et al. Characterization of dengue virus serotype 2 cosmopolitan genotype circulating in colombia. Am J Trop Med Hyg. 2023 Dec 6;109(6):1298–302.

15. Perez-Restrepo LS, Ciuoderis K, Usuga J, Moreno I, Vargas V, Arévalo-Arbelaez Aj, et al. Mayaro Virus as the cause of Acute Febrile Illness in the Colombian Amazon Basin. Front Microbiol. 2024 Jul 9;15:1419637.

