## Appendix for "Co-circulation of two lineages of Oropouche virus in the Amazon basin, Colombia, 2024"

**Table 1.** Protocols of microbiology assays performed at the One Health Colombia, 2024.

| Assay | Pathogen | Protocol |
|---|---|---|
| Oropouche real-time RT-qPCR | OROV | Ciuoderis, et al. Emerg. Microb & Infec. 2022 11:1, 2645-2657 |
| ZCD Trioplex real-time RT-qPCR | ZIKV, CHIKV, DENV | Waggoner, et al. Emerg Infec Dis. 2016 22(7):1295-1297 |
| Mayaro real-time RT-qPCR | MAYV | Gomes Naveca, et al. Mem Inst Oswaldo Cruz . 2017 Jul;112(7):510-513 |
| DENV non-structural protein 1 | DENV | Abbott RDT |
| Anti-DENV IgM | DENV | Abbott RDT |
| Anti -DENV IgG | DENV | Abbott RDT |
| Hepatitis B virus surface Ag | Hepatitis B virus | Abbott Architect |
| Anti-hepatitis C virus antibody | Hepatitis C virus | Abbott Architect |
| Malaria qPCR | *Plasmodium* spp. | Kamau, et al. PLoS One. 2013 29;8(8):e71539 |
| Malaria antigen | *P. vivax* | Abbott RDT |
| Leptospira qPCR | *Leptospira* spp. | Waggoner et al. J Clin Microbiol. 2014 Jun;52(6):2011-8 |

OROV, Oropouche virus; ZIKV, Zika virus; CHIKV, Chikungunya virus; MAYV, Mayaro virus; RDT, rapid diagnostic test.

**Table 2.** Clinical manifestations of the OROV cases in Leticia municipality, Colombia, 2024.

| | $OROV_{PE\text{-}CO\text{-}EC/2008\text{-}2021}$ N=6 (No.) | $OROV_{BR\text{-}2015\text{-}2023}$ N=2 (No.) |
|---|---|---|
| Fever | 1 | 1 |
| Headache | 6 | 2 |
| Myalgia | 5 | 2 |
| Arthralgia | 6 | 1 |
| Chills | 3 | 0 |
| Fatigue | 5 | 2 |
| Rash | 1 | 0 |
| Diarrhea | 3 | 1 |

OROV, Oropouche virus.

**RdRp (L)**

| | 201 | 215 | 244 | 263 | 269 | 303 | 338 | 382 | 429 | 466 | 528 | 565 | 663 | 677 | 739 | 794 | 851 | 940 | 1028 | 1035 | 1114 | 1192 | 1439 | 1758 | 1780 | 1942 | 2116 | 2187 | 2228 | 2230 | 2241 |
|---|---|---|---|---|---|---|---|---|---|---|---|---|---|---|---|---|---|---|---|---|---|---|---|---|---|---|---|---|---|---|---|
| PE/CO/EC 2008-2021 | T | S | K | A | K | I | V | V | N | N | I | A | K | S | N | I | K | Y | N | S | L | V | T | I | G | V | A | I | M | S | K |
| BR 2009-2018 | T | A | R | T | R | M | V | I | S | E | V | A | R | S | S | V | R | H | D | N | L | I | A | V | E | V | V | I | I | N | R |
| BR 2018-2023 | A | S | K | A | K | I | I | V | N | E | I | T | R | A | N | I | K | Y | N | S | V | V | T | I | E | I | A | V | M | S | K |

**Gn (M)**

| | 7 | 35 | 40 | 41 | 75 | 82 | 83 | 207 | 228 |
|---|---|---|---|---|---|---|---|---|---|
| PE/CO/EC 2008-2021 | S | A | A | L | N | L | I | I | R |
| BR 2009-2018 | N | V | R | P | S | I | M | V | K |
| BR 2018-2023 | N | V | R | P | S | I | V | V | K |

**Nsm (M)**

| | 304 | 321 | 322 | 324 | 326 | 333 | 334 | 336 | 337 | 338 | 347 | 352 | 356 | 359 | 363 | 366 | 367 | 368 | 386 | 393 | 402 | 413 | 420 | 422 | 428 | 437 | 439 | 443 | 444 | 445 | 449 | 449 |
|---|---|---|---|---|---|---|---|---|---|---|---|---|---|---|---|---|---|---|---|---|---|---|---|---|---|---|---|---|---|---|---|---|
| PE/CO/EC 2008-2021 | V | D | Y | E | A | G | K | I | K | I | L | I | S | I | Y | N | I | L | V | K | D | V | V | R | G | T | I | T | T | A | S | F |
| BR 2009-2018 | I | E | F | K | A | E | R | M | E | V | L | V | F | M | R | S | I | F | L | K | N | I | I | K | S | I | A | A | L | V | T | I |
| BR 2018-2023 | I | E | F | K | T | E | L | M | E | V | F | V | F | M | R | S | T | F | L | R | N | I | I | K | S | I | A | A | L | V | T | I |

**Gc (M)**

| | 540 | 550 | 557 | 559 | 563 | 598 | 603 | 608 | 610 | 619 | 621 | 625 | 630 | 651 | 672 | 684 | 697 | 756 | 762 | 770 | 776 | 777 | 792 | 804 | 820 | 843 | 899 | 924 | 940 | 995 | 1003 | 1046 |
|---|---|---|---|---|---|---|---|---|---|---|---|---|---|---|---|---|---|---|---|---|---|---|---|---|---|---|---|---|---|---|---|---|
| PE/CO/EC 2008-2021 | F | A | S | F | I | S | N | V | V | N | S | T | Q | S | A | N | V | I | S | D | E | G | A | K | T | S | G | V | K | I | V | N |
| BR 2009-2018 | Y | S | A | L | M | T | D | I | I | D | N | S | H | N | T | S | I | T | T | G | D | S | T | E | S | S | N | A | R | V | V | T |
| BR 2018-2023 | Y | S | A | L | M | T | D | I | I | D | N | S | H | N | T | S | I | T | T | G | D | S | T | E | S | T | N | A | R | V | A | T |

**Gc (M)**

| | 1048 | 1055 | 1063 | 1076 | 1083 | 1085 | 1117 | 1118 | 1123 | 1130 | 1133 | 1152 | 1161 | 1168 | 1186 | 1188 | 1204 | 1208 | 1214 | 1229 | 1231 | 1236 | 1262 |
|---|---|---|---|---|---|---|---|---|---|---|---|---|---|---|---|---|---|---|---|---|---|---|---|
| PE/CO/EC 2008-2021 | M | V | V | V | S | N | N | P | I | I | I | V | V | V | D | P | S | I | T | T | N | N | R |
| BR 2009-2018 | I | I | I | I | T | S | E | K | I | I | I | V | I | I | E | P | N | F | N | I | H | D | R |
| BR 2018-2023 | I | I | I | I | T | S | E | K | T | V | V | I | I | I | E | S | N | F | N | I | H | D | K |

**Figure 1.** Amino acid substitutions in OROV lineages. Amino acid changes found in the virus proteins across three major clades of OROV, PE/CO/EC 2008-2021, BR 2009-2018, and the novel BR 2018-2023 lineage, are shown. A total of 31 changes were observed in the RdRp (L), 9 in Gn (M), 32 in Nsm (M), and 55 in Gc (M). No amino acid changes were found in the S segment. The representative strains for each lineage were determined by calculating the consensus sequence from the respective group of sequences within each lineage.
